# Do latitude and ozone concentration predict Covid-2019 cases in 34 countries?

**DOI:** 10.1101/2020.04.09.20060202

**Authors:** Mark M. Alipio

## Abstract

In this paper, I used multivariate linear regression analysis to determine if latitude and ozone concentration predict Covid-2019 cases in 34 countries worldwide. Data pertaining to Covid-2019 cases were extracted from Worldometer. Ozone concentration levels were taken from the open-access database of World Ozone and Ultraviolet Radiation Data Centre (WOUDC). Latitude of specific area where measurement took place was also provided in the database. Preliminary Kendall rank correlation test revealed that Covid-2019 incidence was positively and significantly related to ozone concentration; however, incidence was not significantly related to latitude. Using multivariate linear regression, a statistically significant link between ozone concentration and Covid-2019 incidence in 34 countries was established; however, I found no statistical association between latitude and Covid-2019 incidence refuting previous claims. Prompt health actions should be developed for areas with high ozone concentration in the present and possibly, future outbreaks; however, extensive laboratory analysis should be conducted to further confirm the findings of the study. Nevertheless, the results of this study could serve as a basis for further clinical and large-scale studies.

## Background

It has come to my attention that Covid-2019 trajectory could be explained by temperature, humidity, and latitude.^1,2,3^ However, prediction needs more sound empirical evidence to provide accurate planning strategies for the present and possibly, future outbreaks. Previous works also provided argument pertaining to the possible relationship between latitude dependence of Covid-2019 mortality and Vitamin D deficiency.^4,5^

Vitamin D is strongly affected by ozone variability, since ozone filters Ultraviolet B, an important factor for Vitamin D synthesis.^6^ A possible link between ozone concentration and Covid-2019 incidence, therefore, should be established. In this paper, I used multivariate linear regression analysis to determine if latitude and ozone concentration predict Covid-2019 cases in 34 countries worldwide.

## Methods

Data pertaining to Covid-2019 cases were extracted from Worldometer.^7^ Ozone concentration levels were taken from the open-access database of World Ozone and Ultraviolet Radiation Data Centre (WOUDC).^8^ Ozone concentration was expressed as Dobson unit (DU), the number of molecules of ozone required to generate a 0.01-mm layer of pure ozone at a temperature of 0 °C and a pressure of 1 atm. Unfortunately, data pertaining to ozone concentration were only available in 34 countries. Measurement was recorded by WOUDC on April 2019 and monthly ozone concentration was considered for the analysis. Latitude of specific area where measurement took place was also provided in the database. The complete data of 34 countries were provided in Table 4.

**Table 1.** Descriptive statistics

| Variables | Frequency, N | Percentage, % | Mean |
| --- | --- | --- | --- |
| Overall | 34 | 100.0 |  |
| Latitude, degrees |  |  | 30.9 |
| -60 to -10.001 | 5 | 14.7 |  |
| -10 to 39.999 | 11 | 32.4 |  |
| 40 to 89.999 | 18 | 52.9 |  |
| Ozone concentration, Dobson unit |  |  | 334.2 |
| 240 - 309.9 | 11 | 32.4 |  |
| 310 - 379.9 | 18 | 52.9 |  |
| 380 - 449.9 | 5 | 14.7 |  |
| Covid-2019 cases, N |  |  | 277.4 |
| 0 - 399 | 41 | 65.1 |  |
| 400 - 799 | 16 | 25.4 |  |
| 800 - 1199 | 6 | 9.5 |  |

**Table 2.** Kendall rank correlation analysis

| Variables | 1 | 2 | 3 |
| --- | --- | --- | --- |
| 1. Latitude | - | 0.44** | 0.08 |
| 2. Ozone concentration |  | - | 0.85*** |
| 3. Covid-2019 cases |  |  | - |

Note: \*p<0.05. \*\*\*p<0.001

**Table 3.** Regression analysis summary for latitude and ozone concentration predicting Covid-2019 cases

| Variables | B | $\beta$ | t | p-value |
| --- | --- | --- | --- | --- |
| (Constant) | 17549.230 |  | 4.170 | <0.001 |
| Latitude | 10.234 | 0.120 | 1.127 | 0.268 |
| Ozone concentration | 706.962 | 0.721 | 4.712 | <0.001 |

**Table 4.** Ozone concentration levels of 34 countries included in the analysis

| Countries | Ozone Concentration, Dobson unit (April 2019) |
| --- | --- |
| Algeria | 265.6 |
| Argentina | 274.2 |
| Australia | 269.2 |
| Belgium | 384.9 |
| Brazil | 260 |
| Chile | 268.5 |
| China | 357.3 |
| Egypt | 312 |
| Finland | 362.2 |
| France | 291.6 |
| Germany | 359.1 |
| Greece | 375.7 |
| Greenland | 415.9 |
| India | 285.9 |
| Ireland | 383.4 |
| Italy | 373.4 |
| Japan | 350.3 |
| Lithuania | 367.2 |
| Luxembourg | 354.3 |
| Moldova | 362.3 |
| Netherlands | 386.2 |
| Norway | 364.3 |
| Philippines | 242.8 |
| Poland | 355.4 |
| Romania | 367.6 |
| Slovakia | 348.4 |
| Spain | 364.8 |
| Sweden | 353.3 |
| Taiwan | 289.9 |
| Thailand | 263.5 |
| UK | 387.1 |
| Ukraine | 363.4 |
| USA | 320.3 |
| Vietnam | 283.2 |

Descriptive statistics (mean, frequency, percentage) were used for the categorical and continuous variables. I used Kendall rank correlation test to determine if latitude and ozone concentration are significantly associated with Covid-2019 cases. Multivariate linear regression was used for the prediction of cases based on latitude and ozone concentration predictors. A p-value below 0.05 was considered significant.

## Results and Discussion

Mean latitude of countries used in the analysis was 30.9 degrees, mean ozone concentration was 334.2 Dobson units, and mean Covid-2019 case was 277.4 cases (Table 1). Kendall rank correlation test revealed that Covid-2019 cases were positively and significantly related to ozone concentration (p<0.001); however, cases were not significantly related to latitude (Table 2). This is indicative that the number of Covid-2019 cases in 34 countries increases with ozone concentration level, while latitude has nothing to do with the burgeoning cases.

Multivariate linear regression revealed that only ozone concentration significantly predicts Covid-2019 cases (β=0.721, p<0.001). For every one Dobson unit increase in the ozone concentration, the number of Covid-2019 cases also increases by approximately more than 706 cases (B=706.962). This shows how an incremental change in ozone concentration could drastically influence the number of Covid-2019 cases across 34 countries studied.

Ozone concentration is inversely related to Ultraviolet transmission.^6^ While other factors may play an important role in Vitamin D synthesis in the human body, it has been recommended that the most promising means of mitigating the burden of Vitamin D deficiency seems to be by increased Ultraviolet exposure (at optimal level).^6^ This highly suggests that high ozone concentration could reduce transmission of Ultraviolet rays, which could have aided synthesis of Vitamin D. Although no clinical trials have been made in the 34 countries included in the study, Vitamin D levels in individuals residing in areas where ozone concentration was high could possibly be deficient or at least insufficient. Studies established how Vitamin D supplementation could increase immunity against infection, especially the common cold.^9,10^

While there is a dearth of research confirming the effect of Vitamin D supplementation on Covid-2019 outcome among actual patients, a previous work found Vitamin D supplementation could possibly improve clinical outcomes of Covid-2019 patients based on increasing odds ratio of having a mild outcome when serum 25-hydroxyvitamin D [25(OH)D] level increases.^5^ Serum 25(OH)D is a robust indicator of Vitamin D status. Hence, health action plans could be developed to provide adequate intake of Vitamin D to individuals living in areas where ozone concentration is high. However, the negative spillover effects of low ozone concentration should not be downplayed as this is an important indicator of ozone layer depletion in the stratosphere.

This study noted several limitations. The sample size is small; hence, generalizability of the results is limited to the countries included in the analysis. Also, actual patients’ information were not used to support the claim on the relationship between Vitamin D supplementation and Covid-2019 clinical outcome. Therefore, clinical trials and large sample studies should be conducted to further verify the causal link among these variables.

## Conclusion

In conclusion, a statistically significant link between ozone concentration and Covid-2019 cases in 34 countries was established in the study; however, I found no statistical association between latitude and Covid-2019 incidence refuting previous claims. Prompt health actions should be developed for areas with high ozone concentration in the present and possibly, future outbreaks; however, extensive laboratory analysis should be conducted to confirm the link between Vitamin D supplementation and Covid-2019 clinical outcome. Nevertheless, the results of this study could serve as a basis for further clinical and large-scale studies.

## Data Availability

The author declares that all data supporting the findings of this study are available within the article and its supplementary information files.

## Supplementary Data

